# Comorbidity trajectories before and after the diagnosis of heart failure: a UK Biobank cohort study

**DOI:** 10.1101/2024.08.28.24312712

**Authors:** Hugo MacGowan, Oliver I Brown, Michael Drozd, Andrew MN Walker, Marilena Giannoudi, Sam Straw, Maria F Paton, John Gierula, Melanie McGinlay, Kathryn J Griffin, Klaus K Witte, Mark T Kearney, Richard M Cubbon

**Affiliations:** Leeds Institute of Cardiovascular and Metabolic Medicine, University of Leeds, UK

**Keywords:** Multimorbidity, Multiple long-term conditions, Comorbidity, Heart failure

## Abstract

**Background:** Heart Failure (HF) is frequently associated with multiple comorbidities. We aimed to define their trajectory of accrual to identify opportunities for disease prevention.

**Methods:** We identified all participants in the UK Biobank cohort study diagnosed with HF prior to enrolment or during follow-up, who had disease occurrence data available from both primary and secondary care records (n=9,824). We established the time between diagnosis of HF and 16 common comorbidities to determine the rate and sequence of comorbidity accrual in relation to HF. Stratified analyses considered associations with sex and age at diagnosis of HF.

**Findings:** In chronological sequence, HF was the median fourth diagnosis for men and women. As the age at HF diagnosis increased, HF came later in the sequence of diseases (median second in under 50s to fifth in those aged 80-90). In all age strata, comorbidities accumulated for over a decade before HF and this accelerated in the years immediately before HF. The median time between comorbidity and HF diagnoses ranged from depression preceding HF by 10.7 years to dementia proceeding HF by 0.7 years; all comorbidities presented earlier in women. Atrial fibrillation/flutter was the commonest disease to immediately precede HF, followed by hypertension, cancer, myocardial infarction and osteoarthritis.

**Interpretation:** HF is most often diagnosed in people with established multimorbidity. There is a protracted window of opportunity during which interventions to prevent HF could be applied, often in disease contexts where this is not routine care, such as cancer and osteoarthritis.

**Funding:** British Heart Foundation (RG/F/22/110076)

## Introduction

An ageing society and improving medical and public health practice mean that a growing proportion of people live with multiple long-term conditions (MLTC, also known as multimorbidity), which can be defined as two or more medical conditions.^1^ A seminal cross-sectional analysis of 1.7 million people living in Scotland in 2007 revealed that a majority of people had MLTC by the age of 65-69.^2^ By their nature, long-term conditions accumulate over time, imposing an increasing burden on individuals, although there is substantial inter-individual heterogeneity in the rate of accrual and the contributing conditions.^3,4^ People with a larger number of medical conditions experience poorer quality of life and a greater burden of medical therapy; they also require substantially larger quantity healthcare resources, such as planned and emergency care contacts.^5^

Chronic heart failure (HF) is a common late phase in the natural history of many cardiovascular diseases, affecting millions of people globally.^6^ Improving treatment over recent decades has been associated with improving survival rates and increasing contribution of non-cardiovascular events to mortality in people with HF.^7^ This has been paralleled by an increasing prevalence of MLTC, such that people recently diagnosed with HF had a median of 5 other long-term conditions (from a possible 17 studied) in a large cross-sectional analysis conducted in England in 2014.^8^ People with HF and a greater number of comorbidities experience greater loss of life expectancy, increased rates of both cardiovascular and non-cardiovascular death, and less commonly achieve optimal HF medical therapy.^9,10^ Hence, prevention of HF in people with MLTC, and *vice versa*, is an important treatment goal. However, little is known about how and when diseases accrue in the lifecourse of people who develop HF, which hinders the targeting of preventative interventions to the right people at the right time. Therefore, we set out to define the trajectory of comorbidity development in people with HF in the UK Biobank (UKB) cohort study, with the goal of understanding preventative opportunities.

## Methods

### Study cohort

UKB is a prospective observational cohort study of 502,462 participants aged 37-73 years, recruited from 22 assessment centres across the United Kingdom (UK) between 2006-10. It is an open access resource developed using UK Government and biomedical research charity funding, which set out to link wide-ranging phenotypic and healthcare record data. Further details of its design and conduct are available at https://www.ukbiobank.ac.uk and our prior publications.^11,12^ UKB received ethical approval from the NHS Research Ethics Service (11/NW/0382); we conducted this analysis under application number 117090. All participants provided written informed consent and the research was conducted in line with the Declaration of Helsinki.

We excluded all participants without record-level access to clinical events documented in primary care (UKB field ID 42040) since these are known to include more complete and timely records than hospital episode statistics (HES) or death registry data.^13^ Within the subgroup of UKB with primary care record data (n=229,944), we then included only those participants with a diagnosis of HF (from primary care, HES or death registry data) at any time in life until the study censorship date of 24^th^ April 2024. This resulted in a cohort of 9,824 participants who developed HF. We applied no other inclusion or exclusion criteria.

### Definitions of HF and comorbidities

All UKB field IDs and disease codes pertaining to our definition of HF, comorbidities and other participant characteristics are presented in **Supplemental Table 1**. We focussed on the comorbid conditions included by Conrad *et al* in their analysis of MLTC in HF,^8^ since this allows comparison of our data with the most comprehensive published study of this phenomenon in the United Kingdom. They included 17 common long-term conditions, of which we excluded dyslipidaemia given that defining its onset is challenging and not all MLTC studies consider this as a primary disease (as opposed to a risk factor). For the remaining 16 conditions, the definitions applied by Conrad *et al* were aligned with disease first occurrence data available through UKB. Where possible, we used UKB-defined first disease occurrence fields to aid reproducibility of our disease definitions; these included: obesity, chronic renal failure, atrial fibrillation/flutter, primary hypertension, myocardial infarction (either non-ST elevation or ST-elevation), asthma, stroke, chronic obstructive pulmonary disease (COPD), dementia (of any cause), anaemia (of any cause), depression, thyroid disease (of any cause), diabetes (except pregnancy-associated), peripheral arterial disease (which includes aortic aneurysm) and cancer (of any type in the UKB cancer registry data). For osteoarthritis, no suitable collated UKB field ID or IDs were available, so we collated relevant read v2 and CTV3 codes from primary care records (Field ID 42040) and ICD9/ICD10 codes from hospital records (UKB Category 2006).

The first occurrence of any comorbidity was included when recorded either through primary care, HES, death registry or cancer registry data; self-reported occurrences were excluded, as were occurrences with invalid dates (prior to birth or in the future). For each comorbidity, we calculated the difference between its date of first diagnosis and the date of first HF diagnosis in order to understand the sequence and relative timing of comorbidity occurrence. To stratify the cohort by sex, we used UKB field ID 31. To stratify the cohort by age at HF diagnosis, we calculated age at HF diagnosis using the date of HF diagnosis in relation to the year and month of birth (UKB field IDs 34 and 52), deriving the following age at HF diagnosis strata: <50 years, 50-60 years, 60-70 years, 70-80 years and 80-90 years.

### Statistical methods

We analysed UKB via the Research Access Platform secure cloud server (https://ukbiobank.dnanexus.com), using RStudio version 4.1.1. Analysis used the R suite ‘tidyverse’,^14^ with plots were compiled using the embedded ‘ggplot2’ package, or the ‘ggridges’ (https://doi.org/10.32614/CRAN.package.ggridges) and ‘networkD3’ (https://doi.org/10.32614/CRAN.package.networkD3) packages. Continuous data are presented as mean and standard deviation, or median and interquartile range when non-normal. Categorical data are presented as percentage and number. Comparison of data between sexes uses unpaired t-tests for normally distributed continuous data, Mann-Whitney tests for non-normally distributed continuous data, and chi-squared tests for categorical data. Comparisons of continuous data across age at diagnosis of HF strata use Kruskal-Wallis tests. In a sensitivity analysis, we excluded participants diagnosed with HF after 24^th^ April 2019 in order to define how a minimum 5-year observation period influenced the temporal distribution of comorbidity diagnoses in relation to HF diagnosis. All statistical tests were 2-sided and statistical significance was defined as p<0.05, Bonferroni adjusted to p<0.003 when performing tests for each of the 16 long-term conditions assessed.

## Results

Of 501,472 UK Biobank participants, 229,944 had available primary care clinical event records. Within this subgroup, we identified 9,824 people (4.3%) with HF diagnosed up to the date of censorship on 24^th^ April 2024. Their characteristics are illustrated in **Table 1**, showing a mean (standard deviation) age at HF diagnosis of 69.2 (8.5) years and a predominance of men (64%). During the observation period of 743,718 person-years, a total of 38,415 comorbid conditions (i.e. not including HF) were first diagnosed, equating to a median of 4 and mean of 3.91 (2.07) comorbidities per person. There were large differences in the prevalence of the 16 comorbidities we studied, ranging from dementia in 6.5% to atrial fibrillation/flutter in 51.1%.

**Table 1:**
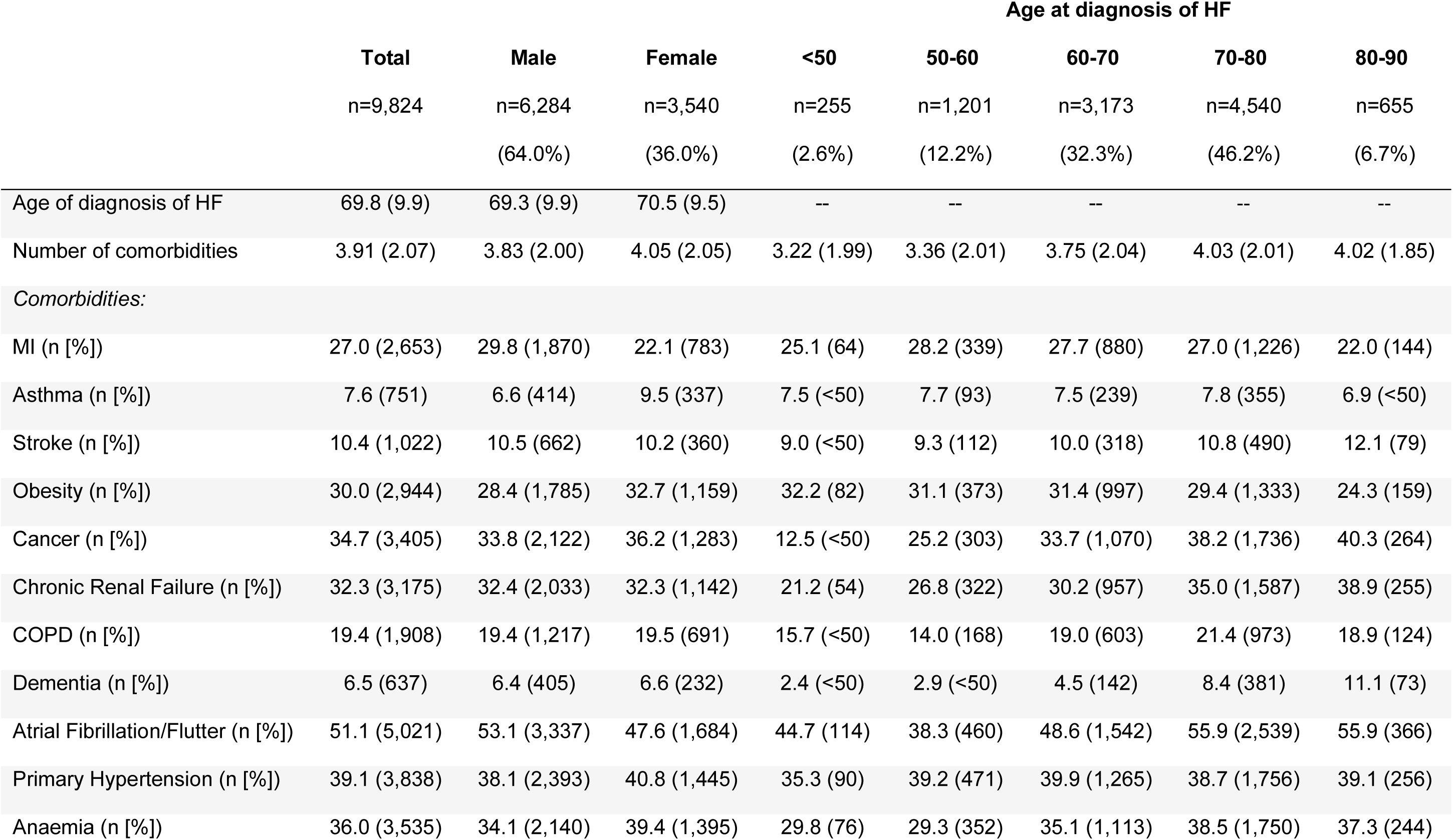
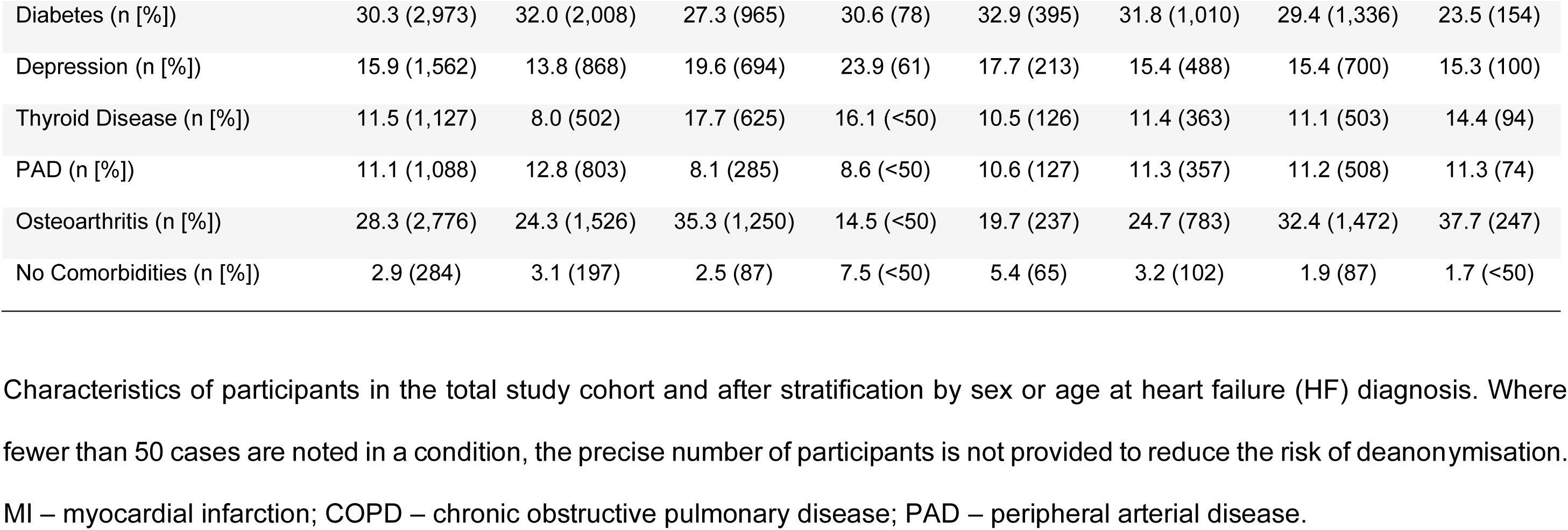
Participant Characteristics.

Men were diagnosed with HF at a younger age than women (68.7 [8.5] versus 70.2 [8.4] years; p<1x10^-10^) and had fewer comorbid conditions (3.83 [2.00] versus 4.05 [2.05] comorbidities per person; p=3.1x10^-5^) during the observation period. Whilst some comorbidities had a similar prevalence in men and women, myocardial infarction, atrial fibrillation/flutter, peripheral arterial disease and diabetes were more common in men, whilst asthma, obesity, anaemia, depression, thyroid disease and osteoarthritis were more common in women (all p<0.003). When stratified according to age at HF diagnosis, there was a small increase in the number of comorbidities from those aged <50 to those aged 70-80, after which this plateaued. Cancer, chronic renal failure, dementia and osteoarthritis showed the largest increase in prevalence with rising age at HF diagnosis, whilst the opposite was noted for obesity, diabetes and depression.

### Sequence and chronology of HF in the trajectory of MLTC accrual

When considered amongst the chronological sequence of MLTC accrual, HF was the median fourth diagnosis received. **Figure 1a** illustrates the distribution of these data, with fewer than 10% of people having HF as their first MLTC diagnosis. When stratified by sex, a subtle rightward shift is noted in women (**Figure 1b**), indicating that HF is a later diagnosis in the sequence of MLTC accrual in women (Mann-Whitney p<1x10^-10^), although HF remains the median fourth diagnosis in men and women. When stratified by age at HF diagnosis, a marked difference in the position of HF in the sequence of MLTC accrual is noted (Kruskal-Wallis p<1x10^-10^; **Figure 1c**). Specifically, HF was the median second diagnosis in people with HF diagnosed under 50 years of age, rising to the median fifth diagnosis in people diagnosed with HF aged 80-90 years of age. Hence, HF is typically diagnosed in the context of established MLTC.

**Figure 1:**
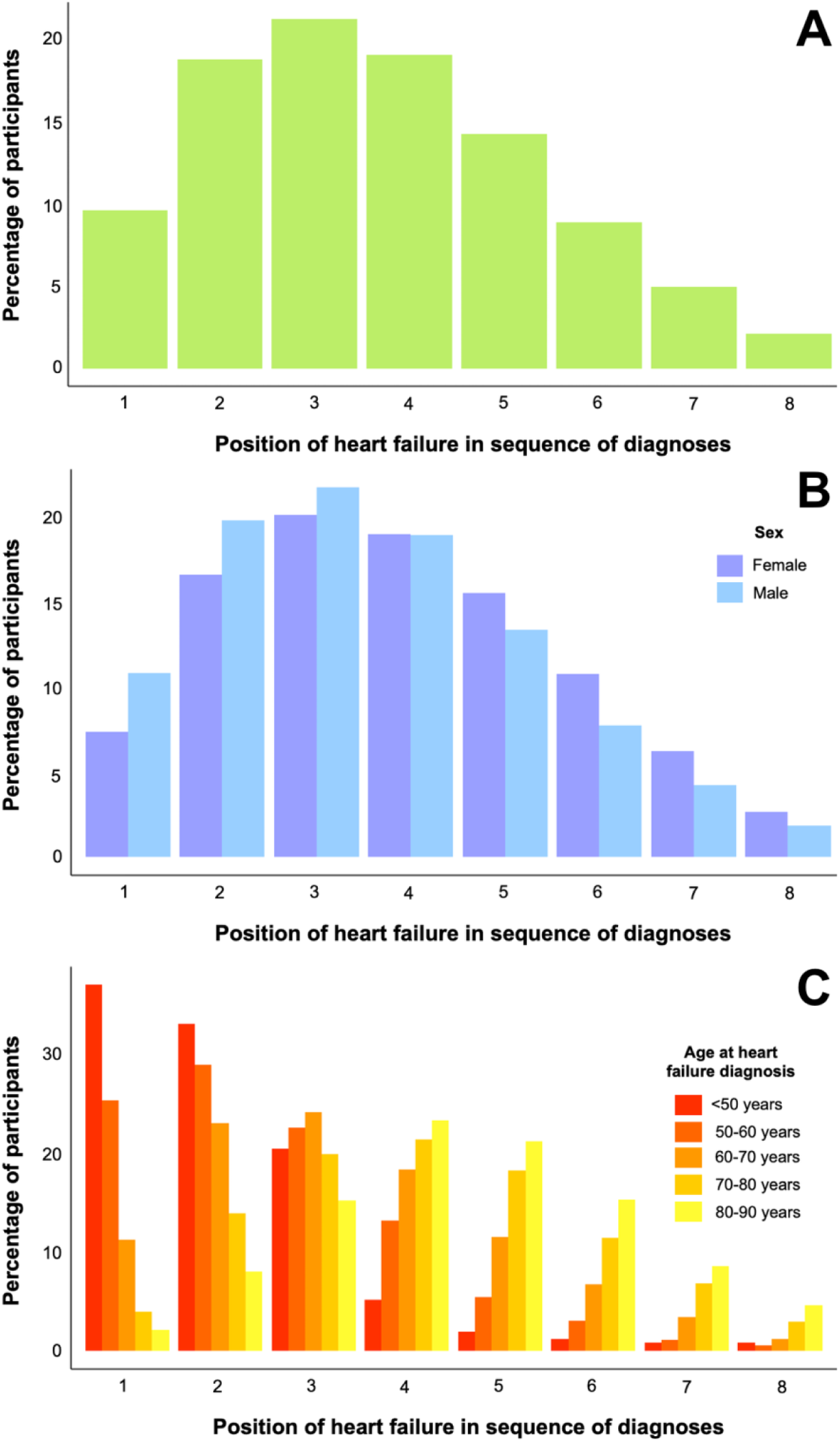
Position of HF in sequence of MLTC diagnoses. Histograms illustrating the position of heart failure (HF) within the sequence of long term condition diagnoses in the whole cohort (**A**), stratified by sex (**B**), or stratified by age at HF diagnosis (**C**).

In order to understand the kinetics of MLTC accrual during the lifecourse of people with HF, we plotted out the number of comorbidities (i.e. excluding HF) over time expressed in relation to the diagnosis of HF (i.e. with negative time representing prior to HF). Notably, data after the diagnosis of HF pertain to a diminishing population as participants die or reach the date of censorship. As people with a greater number of accumulated comorbidities are likely to be older, they are more likely to die which results in a levelling off or reduction in the apparent burden of comorbidities per person. To minimise this phenomenon, we only present data according to strata of age at HF diagnosis (**Figure 2)**. As expected, the number of comorbidities rises over time and is higher in people with HF diagnosed at an older age. However, irrespective of age at HF diagnosis, comorbidities accumulate (at differing rates) for at least 20 years prior to HF diagnosis and accumulate more rapidly around 5 years before HF diagnosis. This period of accelerated comorbidity diagnosis continues for approximately another 5 years after HF diagnosis and then tapers off, especially in those diagnosed with HF later in life. These data illustrate a protracted period of MLTC accrual prior to HF.

**Figure 2:**
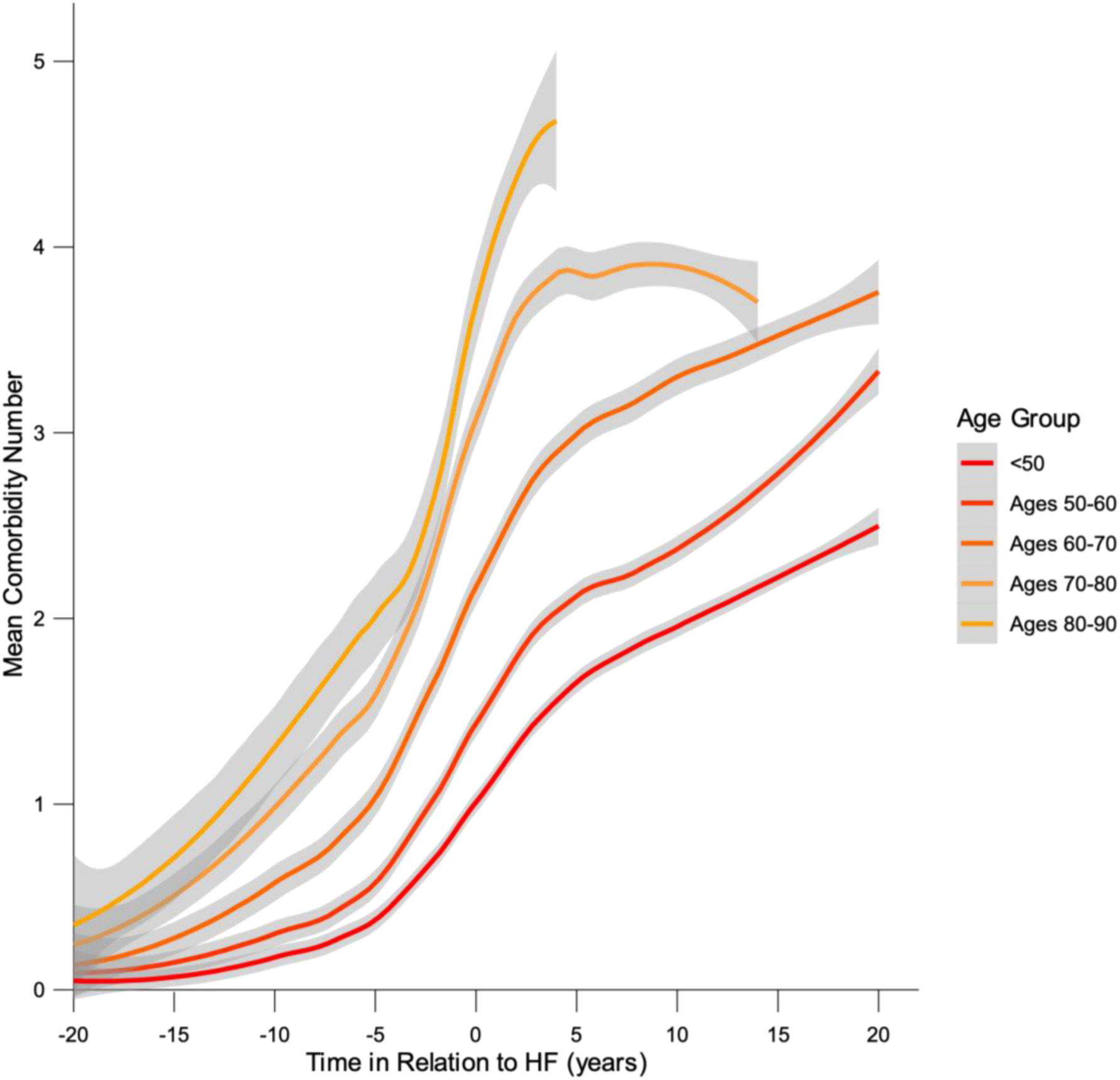
Chronology of comorbidity accrual. For strata at age of heart failure (HF) diagnosis, the mean (and 95% confidence interval of the mean in grey shading) number of non-HF comorbidities are plotted against time relative to HF diagnosis (i.e. negative time denoting prior to HF).

### Timing of individual comorbidity diagnoses in relation to HF

To understand the timing of specific comorbidity diagnoses in relation to HF diagnosis, we constructed Ridgeline plots for each comorbidity (**Figure 3**), again with negative time denoting prior to HF. Importantly, the amplitude of these are not proportional to the prevalence of a comorbidity, allowing a specific focus on the temporal distribution of each comorbidity diagnosis in relation to HF diagnosis. To simplify interpretation, the median and interquartile range is illustrated in each comorbidity Ridgeline (and **Table 2**), and these are ordered according to median time, showing that osteoarthritis is the earliest forerunner of HF and dementia is the latest comorbidity to develop, with a majority of dementia diagnoses occurring after HF. Most cases of some comorbidities present over a narrow period (e.g. myocardial infarction and atrial fibrillation/flutter, with interquartile ranges of 2.4 and 3.9 years, respectively), whereas others present over protracted periods (e.g. depression and cancer, with interquartile ranges of 14.4 and 11.6 years, respectively). For eight of the studied comorbidities (osteoarthritis, depression, obesity, cancer, diabetes, hypertension, atrial fibrillation/flutter and myocardial infarction), three quarters of cases were diagnosed prior to or synchronously with HF. However, it should be noted that with longer follow-up, more cases of these comorbidities would be diagnosed after HF. To address this possibility, we performed a sensitivity analysis including only participants diagnosed with HF on or before 24^th^ April 2019 (n=1,819), giving an opportunity for 5 years of follow-up in those not dying during this period (**Supplemental Table 2**). As expected, in comparison with our primary analysis, this reduced the median time between HF and comorbidity diagnoses for some, with a greater than 1-year difference for thyroid disease, diabetes, cancer, depression and osteoarthritis. Overall, these data emphasise that different comorbidities often have distinct kinetic profiles in relation to the onset of HF.

**Figure 3:**
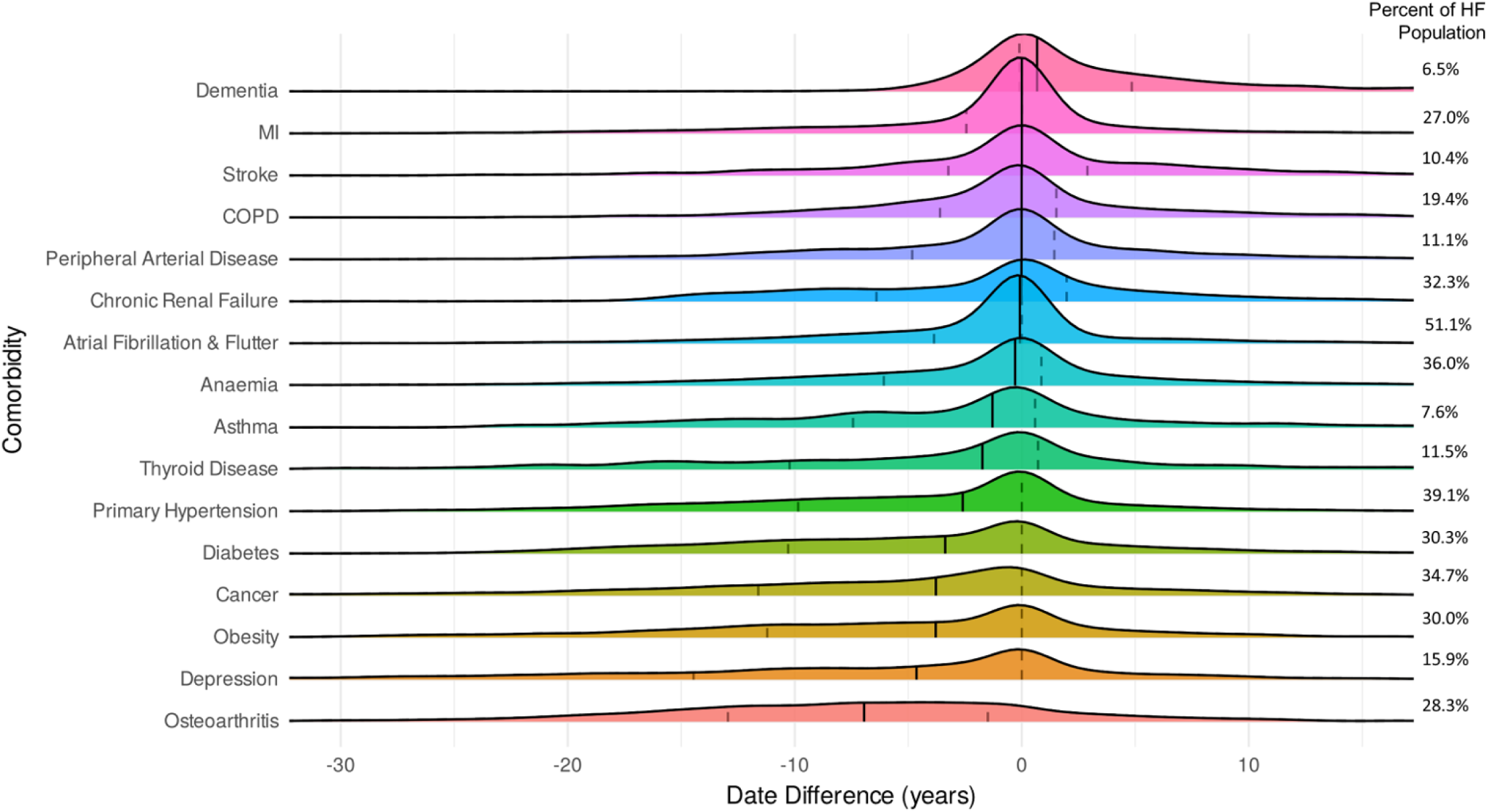
Chronology of comorbidity diagnoses in relation to HF diagnosis. Ridgeline plot illustrating the chronological distribution of comorbidity diagnoses in relation to heart failure (HF) diagnosis. The amplitude of Ridgelines does not relate to comorbidity prevalence, the data for which are noted to the right of the plot for reference. MI - myocardial infarction; COPD – chronic obstructive pulmonary disease.

**Table 2:** Timing of comorbidity diagnoses in relation to HF diagnosis.

| **Comorbidity** | **Time in relation to HF (years)** |
| --- | --- |
| Osteoarthritis | -6.9 (-12.9 to -1.5) |
| Depression | -4.6 (-14.4 to 0.0) |
| Obesity | -3.8 (- 11.2 to 0.0) |
| Cancer | -3.8 (-11.6 to 0.0) |
| Diabetes | -3.4 (-10.3 to 0.0) |
| Primary Hypertension | -2.6 (-9.8 to 0.0) |
| Thyroid Disease | -1.7 (-10.2 to 0.7) |
| Asthma | -1.3 (-7.4 to 0.6) |
| Anaemia | -0.3 (-6.1 to 0.9) |
| Atrial Fibrillation & Flutter | -0.1 (-3.9 to 0.0) |
| Chronic Renal Failure | 0.0 (-6.4 to 2.0) |
| Peripheral Arterial Disease | 0.0 (-4.8 to 1.4) |
| COPD | 0.0 (-3.6 to 1.5) |
| Myocardial Infarction | 0.0 (-2.4 to 0.0) |
| Stroke | 0.0 (-3.2 to 2.9) |
| Dementia | 0.7 (-0.1 to 4.8) |

Data presented as median (first quartile to third quartile)

Next, we explored sex differences in the timing of specific comorbidity diagnoses in relation to HF diagnosis. Ridgeline plots (**Supplemental Figure 1**) and summary data (**Supplemental Table 3**) show that most comorbidities developed earlier, in relation to HF diagnosis, in women than men. This was most pronounced for cancer which developed a median of 7.1 years before HF in women, versus 2.6 years before HF in men. Depression, obesity, thyroid disease and asthma also predated HF by at least 2 years more in women than men. We also generated Ridgeline (**Supplemental Figure 2**) plots and summary data (**Supplemental Table 4**) to address how age at HF diagnosis is associated with the timing of specific comorbidity diagnoses in relation to HF diagnosis. As expected from earlier analyses (**Figure 1c**), in people developing HF before 50 years of age, the majority of cases of each comorbidity developed years after HF (ranging from median of 0 years for myocardial infarction to 13.2 years for dementia). Conversely, in people developing HF aged 80-90 years, the majority of cases of each comorbidity developed years before HF (ranging from median of −11.7 for osteoarthritis to −0.1 years for dementia). Intermediate strata exhibited a spectrum of timings in between these extremes. These data emphasise that sex and age at HF diagnosis are associated with distinct kinetics in the timing of comorbidity development in relation to HF diagnosis.

Finally, we generated a Sankey plot to illustrate the diagnoses occurring immediately before and after HF (**Figure 4**), since the former represent times of opportunity to prevent future HF and the latter opportunities to mitigate comorbidity progression in established HF. Notably, this plot does not illustrate diagnoses coded simultaneously with HF, since these are unlikely to represent new opportunities to prevent disease. However, summary data for this plot in **Table 3** also include synchronous diseases and, as expected, these most often include myocardial infarction (7.3% of HF cases) and atrial fibrillation/flutter (10.9% of HF cases). Atrial fibrillation/flutter is also the most common diagnosis prior to HF (12.9%), with cancer (8.4%), hypertension (7.8%), myocardial infarction (7.0%) and osteoarthritis (7.0%) forming the top 5 prior diagnoses. Regarding diagnoses after HF, these were more evenly distributed, with atrial fibrillation/flutter (6.8%), anaemia (6.3%), chronic renal failure (6.3%), hypertension (4.9%) and cancer (4.4%) forming the 5 commonest diagnoses immediately after HF. These data illustrate both well-established and under-appreciated opportunities for disease prevention, especially before HF.

**Figure 4:**
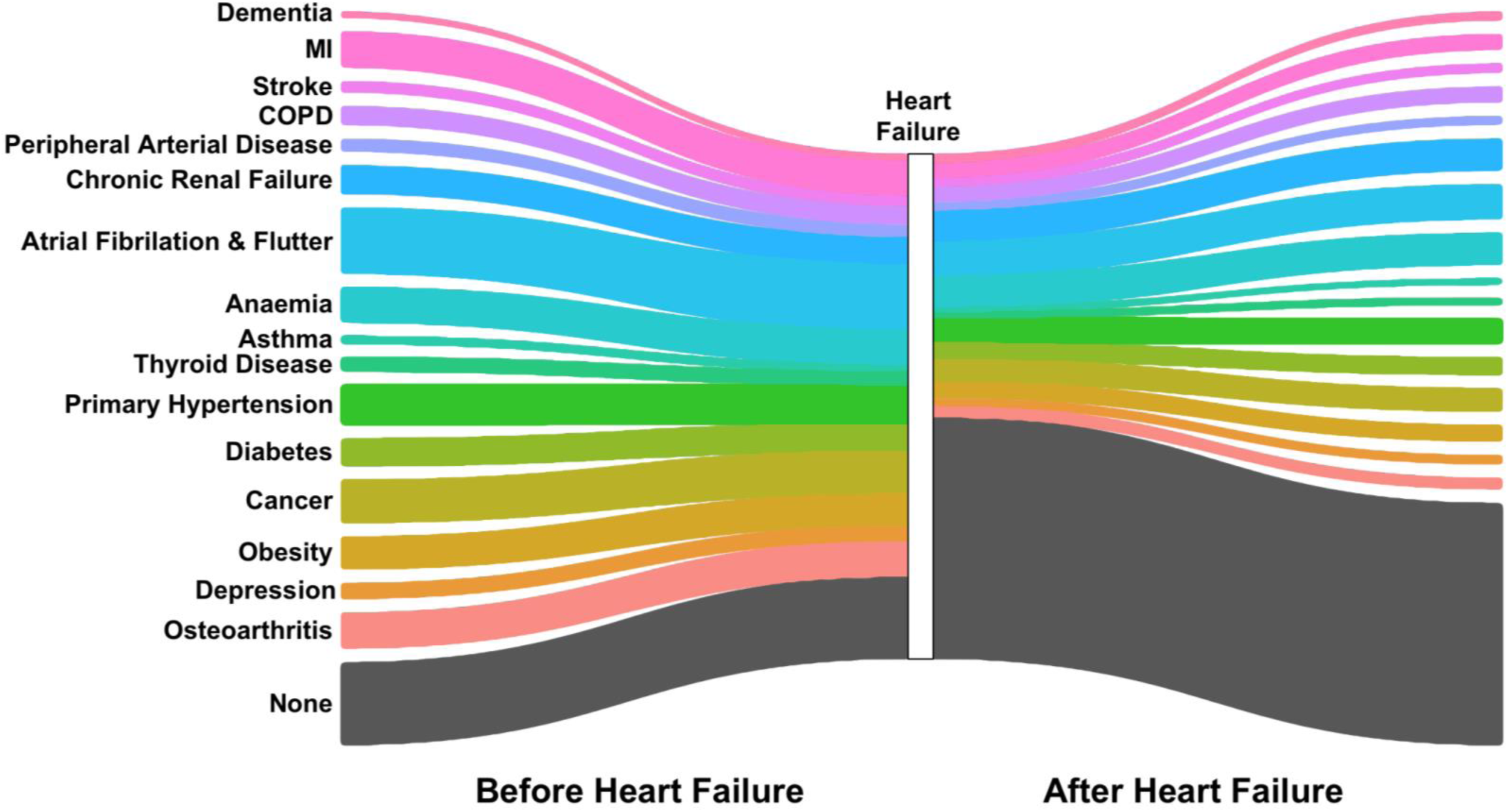
Comorbidity diagnoses immediately before and after HF diagnosis. Sankey plot illustrating the comorbidity immediately before and after heart failure (HF) diagnosis. Comorbidities diagnoses simultaneously with HF are not illustrated (see main text). MI - myocardial infarction; COPD – chronic obstructive pulmonary disease.

**Table 3:** Comorbidities diagnosed immediately before, after or synchronous with HF diagnosis.

| **Comorbidity** | **Immediately before HF** | **Synchronous with HF** | **Immediately after HF** |
| --- | --- | --- | --- |
| Dementia | 1.1 (105) | 0.7 (70) | 1.6 (156) |
| Myocardial Infarction | 7.0 (690) | 7.3 (714) | 3.0 (298) |
| Stroke | 2.1 (204) | 1.0 (102) | 1.7 (168) |
| COPD | 3.6 (358) | 2.7 (263) | 3.0 (299) |
| Peripheral Arterial Disease | 2.4 (240) | 1.2 (121) | 1.6 (159) |
| Chronic Renal Failure | 5.6 (549) | 2.7 (265) | 6.3 (617) |
| Atrial Fibrillation & Flutter | 12.9 (1,268) | 10.9 (1,069) | 6.8 (664) |
| Anaemia | 7.0 (683) | 3.0 (299) | 6.3 (620) |
| Asthma | 1.3 (129) | 0.7 (65) | 0.9 (90) |
| Thyroid Disease | 2.7 (262) | 0.9 (85) | 1.2 (120) |
| Primary Hypertension | 7.8 (762) | 4.2 (408) | 4.9 (482) |
| Diabetes | 5.4 (528) | 2.5 (243) | 3.3 (324) |
| Cancer | 8.4 (824) | 0.4 (<50) | 4.4 (435) |
| Obesity | 6.4 (631) | 2.5 (242) | 3.1 (308) |
| Depression | 3.0 (297) | 0.8 (80) | 1.7 (169) |
| Osteoarthritis | 7.0 (688) | 0.5 (<50) | 2.1 (206) |
| None | 16.3 (1,606) | 6,698 (68.2) | 48.0 (4,714) |

Data presented as percentage (number). Where fewer than 50 cases are noted in a condition, the precise number of participants is not provided to reduce the risk of deanonymisation. COPD – chronic obstructive pulmonary disease.

## Discussion

Our findings have a number of novel and potentially important implications for the prevention and treatment of HF and its comorbidities. First, HF tends to be diagnosed after a sustained period of disease accrual, often spanning over a decade, with an accelerated phase closer to the diagnosis of HF. This suggests that there is a protracted opportunity to address modifiable risk factors for HF in people entering the early phase of multimorbidity. Indeed, we show that hypertension, obesity and diabetes are common antecedents of HF, and it is well established that optimisation of these can reduce incident HF. However, cancer and osteoarthritis were also common precursors of HF, raising the question of whether traditional cardiovascular risk modification could benefit future health in these circumstances. Moreover, understanding why diseases like cancer and osteoarthritis commonly precede HF could lead to additional approaches to reduce incident HF. Our data also highlight opportunities to prevent comorbidity accrual after the diagnosis of HF; in particular, most cases of dementia develop after HF, yet this is not currently considered in the approach to HF care.^15^ Finally, we noted important differences in the patterns of disease accrual between sexes and particularly in relation to the age of HF diagnosis. These should be taken into consideration in strategies to reduce disease accrual, for example prioritising HF prevention in older multimorbid people, versus slowing multimorbidity progression in younger people with HF.

The wider disease context of HF is increasingly important, as evidenced by the large and growing contribution of non-cardiovascular events to hospitalisation and mortality in people with HF.^7,16,17^ Moreover, evidence-based care of HF is harder to achieve and sustain in people with wider health problems.^10,18^ The work of Conrad *et al* has previously shown a significant burden of MLTC in people with HF, rising from three to five non-HF comorbidities per person between 2002 and 2014.^8^ Their work in a large nationally representative sample of the UK population broadly agrees with our finding that multimorbidity is established at the point of HF diagnosis. However, in spite of considering the same comorbidities, our data suggest a slightly lower comorbidity burden at HF diagnosis, probably resulting from our cohort being seven years younger, along with UK Biobank participants being healthier than the UK population.^19^

Our data provide a lifecourse context to what has been shown at the moment of HF diagnosis, thereby identifying potential windows of opportunity for prevention of HF and its comorbidities. At a population level, it is striking that long-term conditions accrued for approximately two decades in advance of HF, irrespective of the age at which HF developed, with the rate of accrual being much faster in older groups. Whilst some individuals developed HF as their first or second major health problem, especially in younger people, the clear majority had a protracted period of multimorbidity, including many non-cardiovascular diseases, prior to the diagnosis of HF. This suggests that interventions to reduce HF risk could be targeted over an extended period of life, often in disease settings (such as osteoarthritis) where current practice would not consider this, or even as part of novel services designed to improve outcomes for people with MLTC.

Whilst simple interventions like optimisation of blood pressure are likely to be useful in many HF prevention settings, addressing the reasons why multiple diseases cluster together may have a larger therapeutic overall impact. One example of this is emerging data on ‘innate immune training’ induced by one disease leading to an inflammatory state that promotes accumulation of other diseases. Indeed, preclinical models show that stroke promotes HF, and that HF promotes kidney disease.^20,21^ Further support comes from *post hoc* analyses of the CANTOS trial, showing that interleukin-1β inhibition reduced incident anaemia, gout, osteoarthritis, lung cancer and heart failure in addition to its primary endpoint of atherosclerotic cardiovascular events.^22–27^ Much greater understanding is still needed from mechanistic, epidemiological and applied health research, but considering diseases in clusters has significant potential to improve the efficacy of treatment and mitigate its burden. However, our data also suggest important associations of age and sex with the nature and rate of MLTC accrual, which will also be important considerations in optimal disease prevention approaches.

Two of the precursors to HF that we identified warrant further discussion. Cancer and osteoarthritis are non-cardiovascular disorders already known to be associated with a higher risk of incident HF, with particularly good evidence for the former.^28,29^ Indeed, there are established clinical guidelines advocating traditional cardiovascular prevention approaches for people receiving the most cardiotoxic cancer therapy,^30^ although adherence to such approaches is low and declines after cancer therapy.^31^ Consideration or modification of future HF risk is not currently included in guidelines on osteoarthritis care. As noted earlier, these complexity of considering multiple single diseases in parallel, rather than as related disease clusters, may underpin these missed opportunities. Changing this approach will require understanding of how these diseases interact with HF, robust clinical evidence that interventions can target multiple diseases, and involvement of patients to develop approaches that are acceptable and minimise treatment burden.

Although our analysis has led to potentially important findings, we must also address its limitations. First, and as alluded to earlier, UK Biobank is not representative of the general UK population, especially in terms of age, ethnicity and socioeconomic deprivation, and exhibits a ‘healthy cohort’ effect.^19^ This means that we may have underestimated the prevalence of MLTC in people with HF, and that our findings may not reliably generalise to the wider UK population or beyond. Second, whist we have lifecourse data in all participants prior to HF, we do not have systematic follow-up for the remainder of the lifecourse, due to participants reaching the censorship date. This biases our analysis of the distribution of incident diseases to the time before HF and also complicates analysis after HF given that the most multimorbid participants are more likely to die, both due to their accumulated disease and older age. Our sensitivity analysis goes some way to addressing this and suggests only a modest effect on our estimates of disease timing in relation to HF, but this should still be carefully considered when using our data. Third, MLTC research is hampered by a lack of consensus on which conditions to consider. Whilst recent attempts have been made to address this,^32^ we elected to replicate definitions used in the most robust available study in HF. Applying other combinations of long term conditions might lead to differing conclusions, but this challenge impacts the entire field of MLTC research. Fourth, misclassification of HF and other long-term conditions in routine care and its subsequent coding is a limitation of healthcare record-based research and may add noise to our dataset. Finally, we do not attempt to subclassify HF or its comorbidities, for example based on the basis of severity, and nor can we account for disease remission. These again reflect a challenge for the field of MLTC research that will require more advanced data handling and analytical approaches. This issue is most pertinent to the classification of HF into that with preserved or reduced left ventricular ejection fraction i.e. HFpEF versus HFrEF. Approximately half of HF is associated with preserved ejection fraction, which is associated with older age, female sex and a greater burden of comorbidities.^33^ Although this would have been an important factor to include in our stratified analyses, these data are not available in UK Biobank, nor UK electronic health records more generally.

## Conclusions

HF is typically diagnosed after a sustained period of disease accrual, often spanning more than a decade, with an accelerated phase closer to the diagnosis of HF. This represents an important window of opportunity for measures to prevent HF, including in the context of diseases, such as cancer and osteoarthritis, where future HF risk is not systematically addressed. Beyond traditional HF prevention approaches, further research is needed to understand the shared basis of these diseases, which may allow the development of interventions addressing multiple conditions.

## Acknowledgements

This research was funded by the British Heart Foundation (RG/F/22/110076).

This research has been conducted using the UK Biobank resource - application number 117090. This work uses data provided by patients and collected by the NHS as part of their care and support. This research used data assets made available by National Safe Haven as part of the Data and Connectivity National Core Study, led by Health Data Research UK in partnership with the Office for National Statistics and funded by UK Research and Innovation (research which commenced between 1st October 2020 – 31st March 2021 grant ref MC_PC_20029; 1st April 2021 -30th September 2022 grant ref MC_PC_20058). HM received a research bursary from The Wolfson Foundation. MD and SS are supported by National Institute for Health and Care Research (NIHR) Academic Clinical Lectureships. MTK is a British Heart Foundation professor. RC receives support from the NIHR Leeds Biomedical Research Centre. The views expressed are those of the author(s) and not necessarily those of the NHS, the NIHR or the Department of Health and Social Care.

## Conflicts of Interest

SS has received speaker’s fees, honoraria and non-financial support from Astra Zeneca. MFP has received consultancy fees from Astra Zeneca. RC has received speaker’s fees from Janssen Oncology.

## Data availability

UK Biobank data are available via application at https://www.ukbiobank.ac.uk

---

**Supplemental Table 1:** UK Biobank definitions of HF and comorbidities.

| **Characteristic** | **Definition Source** |
| --- | --- |
| Anaemia | Field IDs 130622, 130623, 130624, 130625,<br>130626, 130627, 130628, 130629, 130648, 130649,<br>130642, 130643, 130640, 130641, 130636, 130637,<br>130638, 130639, 130634, 130635 |
| Asthma | Field IDs 42014, 42015 |
| Atrial Fibrillation and Flutter | Field IDs 131350, 131351 |
| Cancer | Curated from Field IDs 40009 and 40005 |
| Chronic Renal Failure | Field IDs 132032, 132033 |
| COPD | Field IDs 42016, 42017 |
| Dementia | Field IDs 42018, 42019 |
| Depression | Field IDs 130892, 130893, 130894, 130895,<br>130896, 130897 |
| Diabetes | Field IDs 130706, 130707, 130708, 130709,<br>130710, 130711, 130712, 130713 |
| Heart Failure | Field IDs 131354, 131355 |
| Myocardial Infarction | Field IDs 42000, 42001 |
| Obesity | Field IDs 130792, 130393 |
| Osteoarthritis | Field IDs 42040, 41262, 41202, 41263<br>Secondary care: ICD10 Codes M150-154, M158-M167, M169-M175, M179-M185, M189-M192,<br>M198 and ICD9 Codes 715, 7150, 7151, 7152, |
|  | 7153, 7158, 7159. Primary care Read V2 Code<br>N05.. and Read CTV3 Code XE1DV |
| Peripheral Arterial Disease | Field IDs 131388, 131389, 131390, 131391,<br>131384, 131385, 131382, 131383 |
| Primary Hypertension | Field IDs 131286, 131287 |
| Stroke | Field IDs 42006, 42007 |
| Thyroid Disease | Field IDs 130720, 130721, 130722, 130723,<br>130692, 130693, 130694, 130695, 130696, 130697,<br>130698, 130699, 130700, 130701, 130702, 130703,<br>130704, 130705 |
| Sex | Field ID 31 |
| Month of birth | Field ID 52 |
| Year of birth | Field ID 34 |

Further details relating to these codes are provided by UK Biobank via the online data showcase: https://biobank.ndph.ox.ac.uk/showcase/

**Supplemental Table 2:** Timing of comorbidity diagnoses in relation to HF diagnosis in participants with at least 5 years follow-up or death in this period.

| **Comorbidity** | **Time in relation to HF (years)** |  |
| --- | --- | --- |
|  | **Primary analysis** | **Sensitivity analysis** |
| Osteoarthritis | -6.9 (-12.9 to -1.5) | -5.4 (-11.9 to 0.0) |
| Depression | -4.6 (-14.4 to 0.0) | -2.5 (-11.2 to 0.4) |
| Obesity | -3.8 (- 11.2 to 0.0) | -3.0 (-10.7 to 0.9) |
| Cancer | -3.8 (-11.6 to 0.0) | -2.0 (-9.4 to 1.1) |
| Diabetes | -3.4 (-10.3 to 0.0) | -2.1 (-9.5 to 1.5) |
| Primary Hypertension | -2.6 (-9.8 to 0.0) | -1.6 (-8.8 to 0.5) |
| Thyroid Disease | -1.7 (-10.2 to 0.7) | -0.6 (-9.4 to 2.3) |
| Asthma | -1.3 (-7.4 to 0.6) | -0.5 (-6.8 to 3.3) |
| Anaemia | -0.3 (-6.1 to 0.9) | -0.1 (-5.2 to 1.8) |
| Atrial Fibrillation & Flutter | -0.1 (-3.9 to 0.0) | 0.0 (-3.2 to 0.3) |
| Chronic Renal Failure | 0.0 (-6.4 to 2.0) | 0.0 (-4.5 to 3.5) |
| Peripheral Arterial Disease | 0.0 (-4.8 to 1.4) | 0.0 (-4.5 to 2.5) |
| COPD | 0.0 (-3.6 to 1.5) | 0.0 (-2.7 to 3.4) |
| Myocardial Infarction | 0.0 (-2.4 to 0.0) | 0.0 (-1.6 to 0.1) |
| Stroke | 0.0 (-3.2 to 2.9) | 0.0 (-2.0 to 4.7) |
| Dementia | 0.7 (-0.1 to 4.8) | 0.6 (-0.1 to 5.5) |

Data presented as median (first quartile to third quartile).

**Supplemental Table 3:** Timing of comorbidity diagnoses in relation to HF diagnosis stratified by sex.

| **Comorbidity** | **Time in relation to HF (years)** |  |
| --- | --- | --- |
|  | **Male** | **Female** |
| Osteoarthritis | -6.2 (-12.5 to -0.9) | -7.5 (-13.4 to -2.1) |
| Depression | -3.6 (-13.4 to 0.1) | -6.4 (-15.7 to 0.0) |
| Obesity | -3.0 (-10.5 to 0.2) | -5.3 (12.2 to 0.0) |
| Cancer | -2.6 (-9.3 to 0.8) | -7.1 (-15.6 to -0.8) |
| Diabetes | -3.3 (-10.0 to 0.0) | -3.4 (-10.9 to 0.0) |
| Primary Hypertension | -2.0 (-9.5 to 0.0) | -3.4 (-10.4 to 0.0) |
| Thyroid Disease | 0.0 (-5.8 to 2.3) | -4.1 (-13.7 to 0.0) |
| Asthma | -0.3 (-5.6 to 2.0) | -2.9 (-10.1 to 0.0) |
| Anaemia | 0.0 (-4.5 to 2.1) | -1.5 (-8.6 to 0.2) |
| Atrial Fibrillation & Flutter | -0.1 (-3.9 to 0.0) | -0.1 (-3.8 to 0.0) |
| Chronic Renal Failure | 0.0 (-5.7 to 2.4) | -0.5 (-7.3 to 1.3) |
| Peripheral Arterial Disease | 0.0 (-4.8 to 1.9) | -0.1 (-4.9 to 0.6) |
| COPD | 0.0 (-2.7 to 2.1) | -0.3 (-4.8 to 0.6) |
| Myocardial Infarction | 0.0 (-3.0 to 0.0) | 0.0 (-1.6 to 0.0) |
| Stroke | 0.0 (-3.3 to 3.9) | 0.0 (-2.9 to 1.7) |
| Dementia | 0.7 (-0.1 to 4.6) | 0.0 (-0.2 to 5.1) |

Data presented as median (first quartile to third quartile)

**Supplemental Table 4:** Timing of comorbidity diagnoses in relation to HF diagnosis stratified by age at HF diagnosis.

| Comorbidity | Time in relation to HF (years) |  |  |  |  |
| --- | --- | --- | --- | --- | --- |
|  | HF aged <50 | HF aged 50-60 | HF aged 60-70 | HF aged 70-80 | HF aged 80-90 |
| Osteoarthritis | 7.0 (1.5 to 12.2) | 0.7 (-4.7 to 7.9) | -3.8 (-9.6 to 0.3) | -8.8 (-14.4 to -3.8) | -11.7 (-16.3 to -6.8) |
| Depression | 0.5 (-5.0 to 7.0) | -1.1 (-8.5 to 3.7) | -4.3 (-13.4 to 0.1) | -6.5 (-17.2 to -0.1) | -7.6 (-18.6 to -0.7) |
| Obesity | 0.8 (-2.9 to 9.1) | 0.0 (-5.0 to 6.0) | -3.1 (-10.2 to 0.4) | -5.7 (-13.0 to 0.0) | -8.3 (-14.8 to -2.1) |
| Cancer | 0.6 (-7.8 to 11.7) | 1.9 (-4.4 to 9.0) | -1.2 (-8.3 to 2.9) | -5.8 (-13.0 to -0.9) | -10.3 (-16.9 to -4.0) |
| Diabetes | 5.5 (0.0 to 13.6) | 0.0 (-4.2 to 6.4) | -2.3 (-8.6 to 0.5) | -5.8 (-12.4 to -0.3) | -7.9 (-14.8 to -2.3) |
| Primary Hypertension | 1.4 (-0.1 to 10.0) | 0.0 (-3.0 to 4.1) | -0.4 (-7.4 to 0.9) | -5.3 (-12.3 to -0.1) | -8.1 (-13.4 to -2.2) |
| Thyroid Disease | 8.2 (0.5 to 14.1) | 1.3 (-2.8 to 6.9) | -0.4 (-7.8 to 1.6) | -4.0 (-13.8 to 0.0) | -3.9 (-14.2 to -0.5) |
| Asthma | 1.7 (-1.9 to 13.0) | 0.0 (-1.8 to 10.9) | -0.1 (-6.2 to 2.3) | -3.1 (-8.6 to 0.0) | -6.8 (-12.1 to -0.5) |
| Anaemia | 7.9 (0.0 to 16.2) | 0.6 (-1.9 to 8.1) | 0.0 (-4.8 to 2.9) | -1.0 (-6.9 to 0.1) | -2.6 (-8.9 to 0.0) |
| Atrial Fibrillation & Flutter | 4.9 (0.0 to 13.1) | 0.0 (-0.3 to 6.5) | 0.0 (-2.2 to 0.1) | -0.4 (-5.3 to 0.0) | -2.0 (-7.7 to 0.0) |
| Chronic Renal Failure | 8.3 (0.4 to 15.8) | 4.8 (0.0 to 10.1) | 0.3 (-2.9 to 4.2) | -1.4 (-8.2 to 0.2) | -5.2 (-12.6 to 0.0) |
| Peripheral Arterial Disease | 2.9 (0.0 to 14.5) | 2.7 (-0.7 to 9.2) | 0.0 (-2.3 to 4.0) | -1.0 (-6.6 to 0.0) | -2.0 (-7.5 to 0.0) |
| COPD | 8.0 (1.5 to 14.2) | 4.8 (0.0 to 11.4) | 0.0 (-2.1 to 4.2) | -0.6 (-4.8 to 0.0) | -1.8 (-6.3 to 0.0) |
| Myocardial Infarction | 0.0 (-0.4 to 3.5) | 0.0 (-0.6 to 0.0) | 0.0 (-1.6 to 0.1) | 0.0 (-3.4 to 0.0) | -1.3 (-6.4 to 0.0) |
| Stroke | 8.3 (4.6 to 16.0) | 4.7 (0.0 to 11.3) | 0.2 (-0.9 to 5.8) | -0.2 (-4.6 to 0.7) | -0.8 (-7.3 to 0.0) |
| Dementia | 13.2 (9.2 to 18.6) | 15.9 (10.4 to 18.8) | 6.0 (0.7 to 10.1) | 0.3 (-0.3 to 2.9) | -0.1 (-2.0 to 0.0) |

Data presented as median (first quartile to third quartile)

**Supplemental Figure 1:**
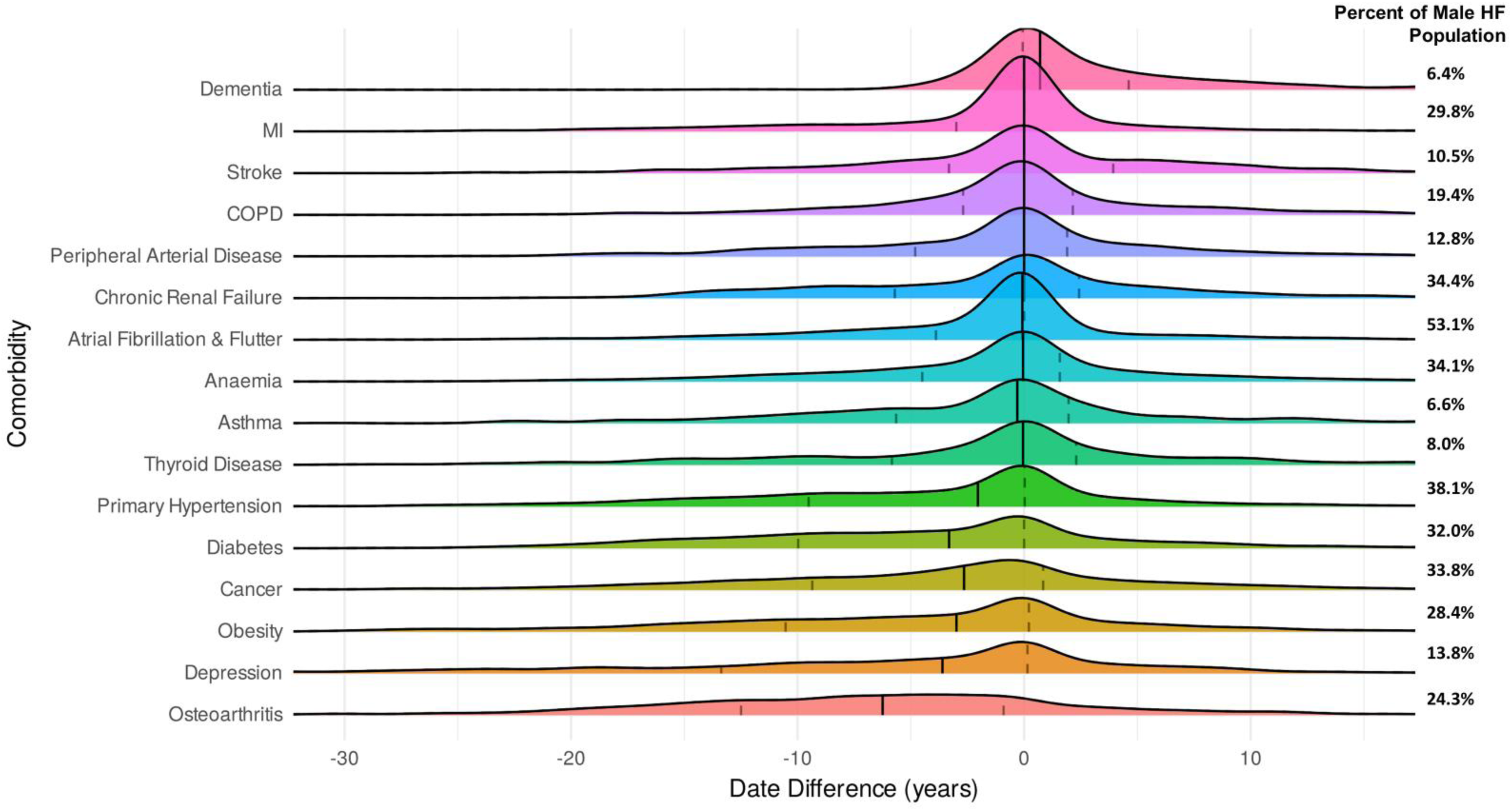
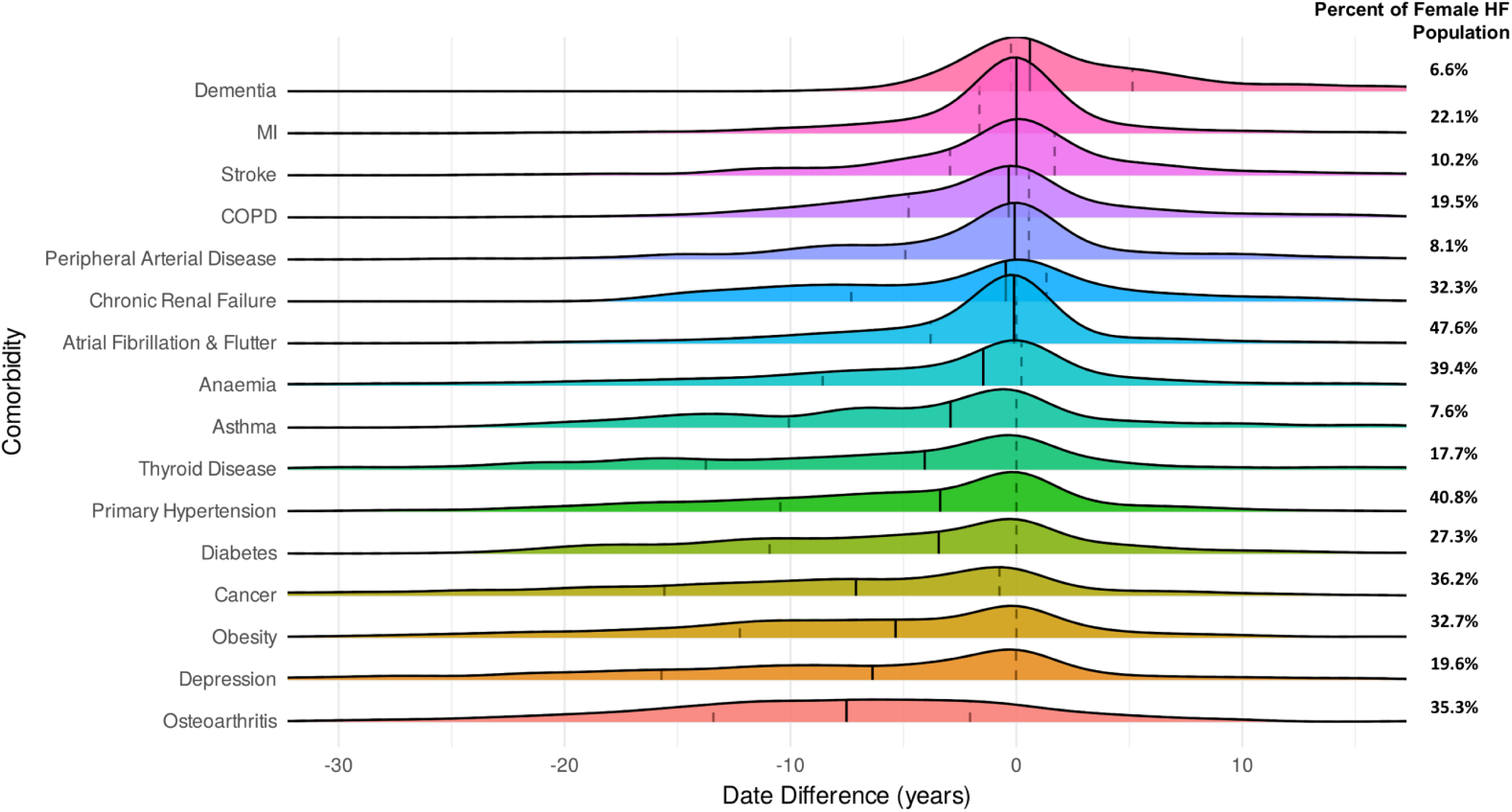
Timing of comorbidity diagnoses in relation to HF diagnosis stratified by sex. Ridgeline plots illustrating the chronological distribution of comorbidity diagnoses in relation to heart failure (HF) diagno sis in men and women. The amplitude of Ridgelines does not relate to comorbidity prevalence, the data for which are noted to the right of the plot for reference. MI - myocardial infarction; COPD – chronic obstructive pulmonary disease.

**Supplemental Figure 2:**
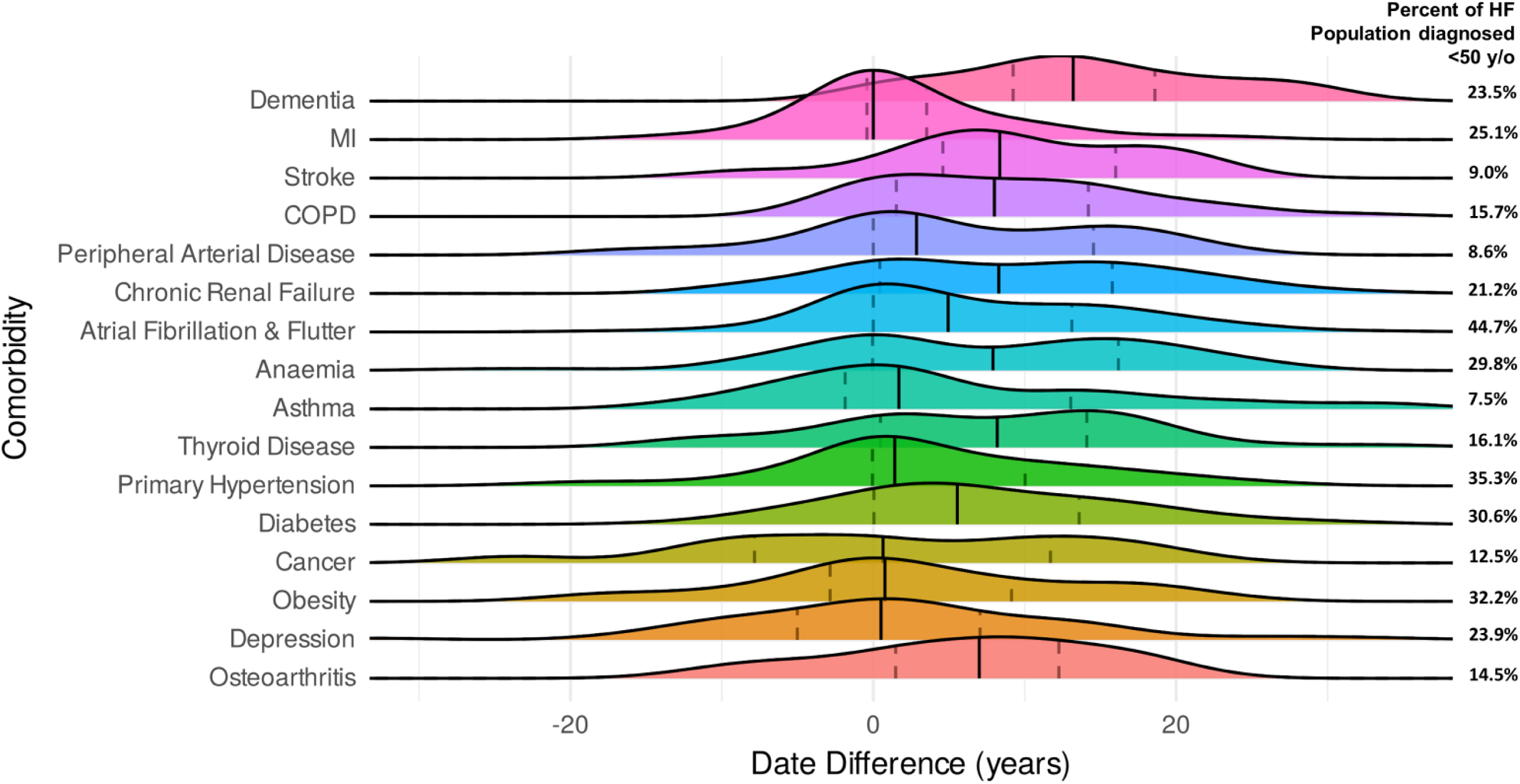
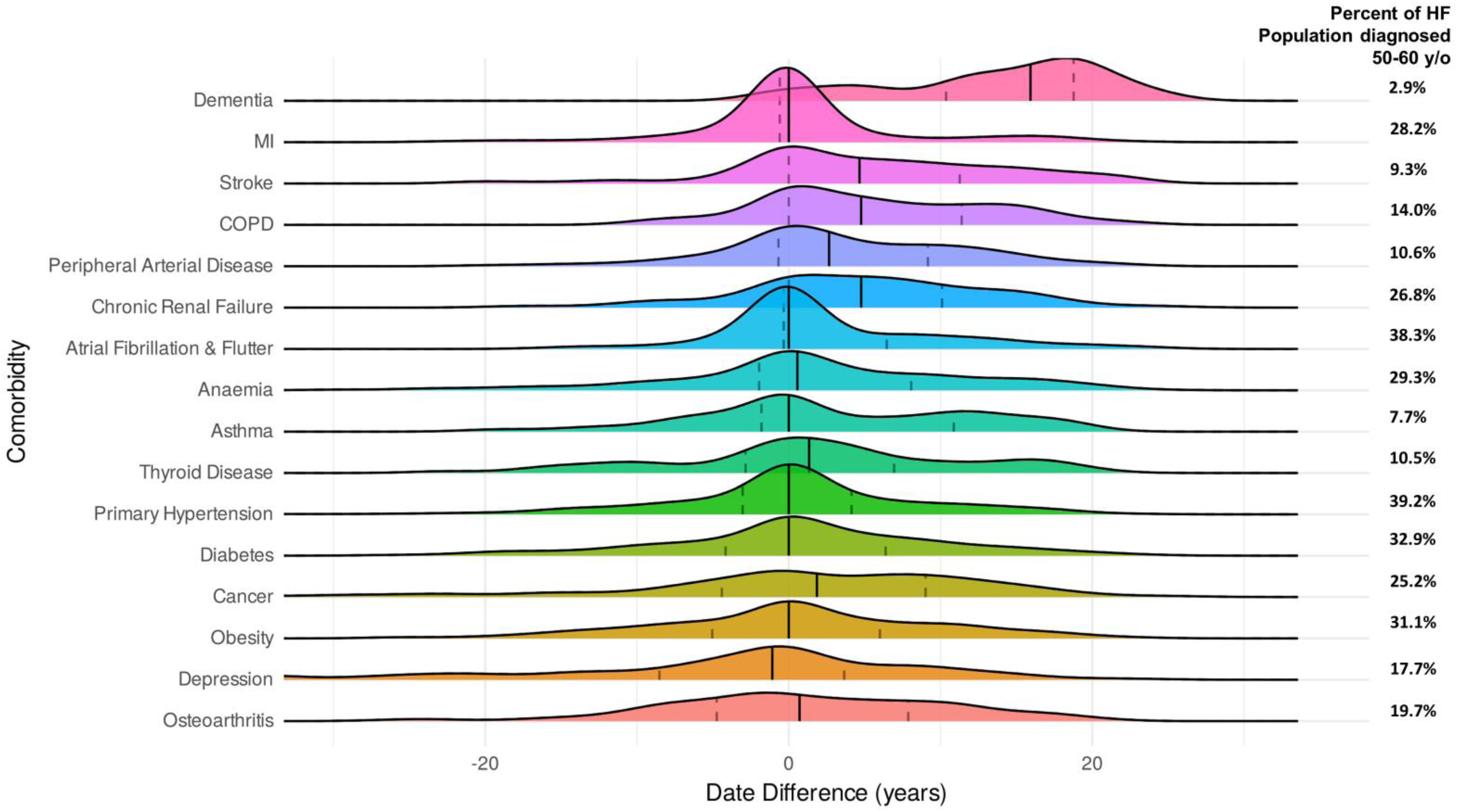
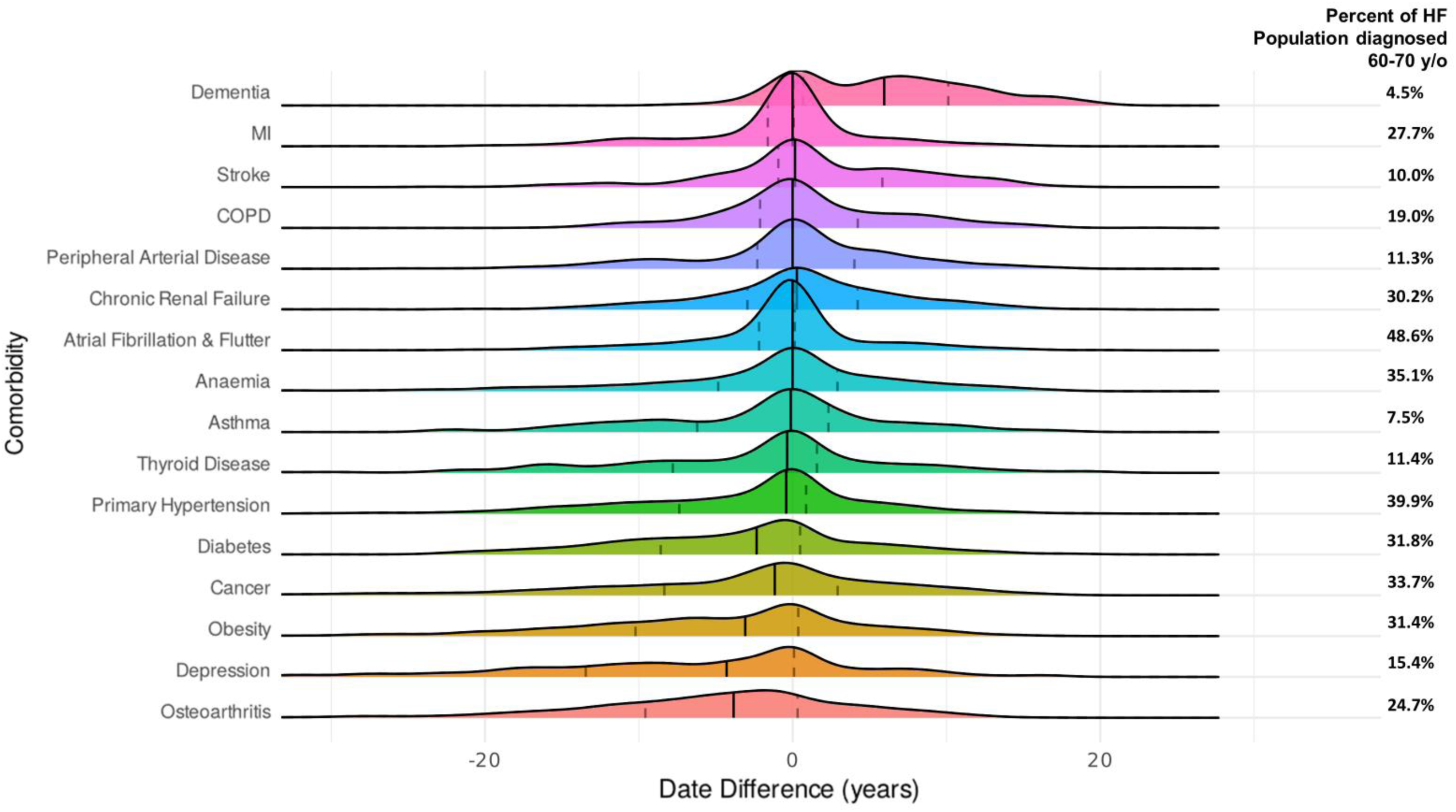
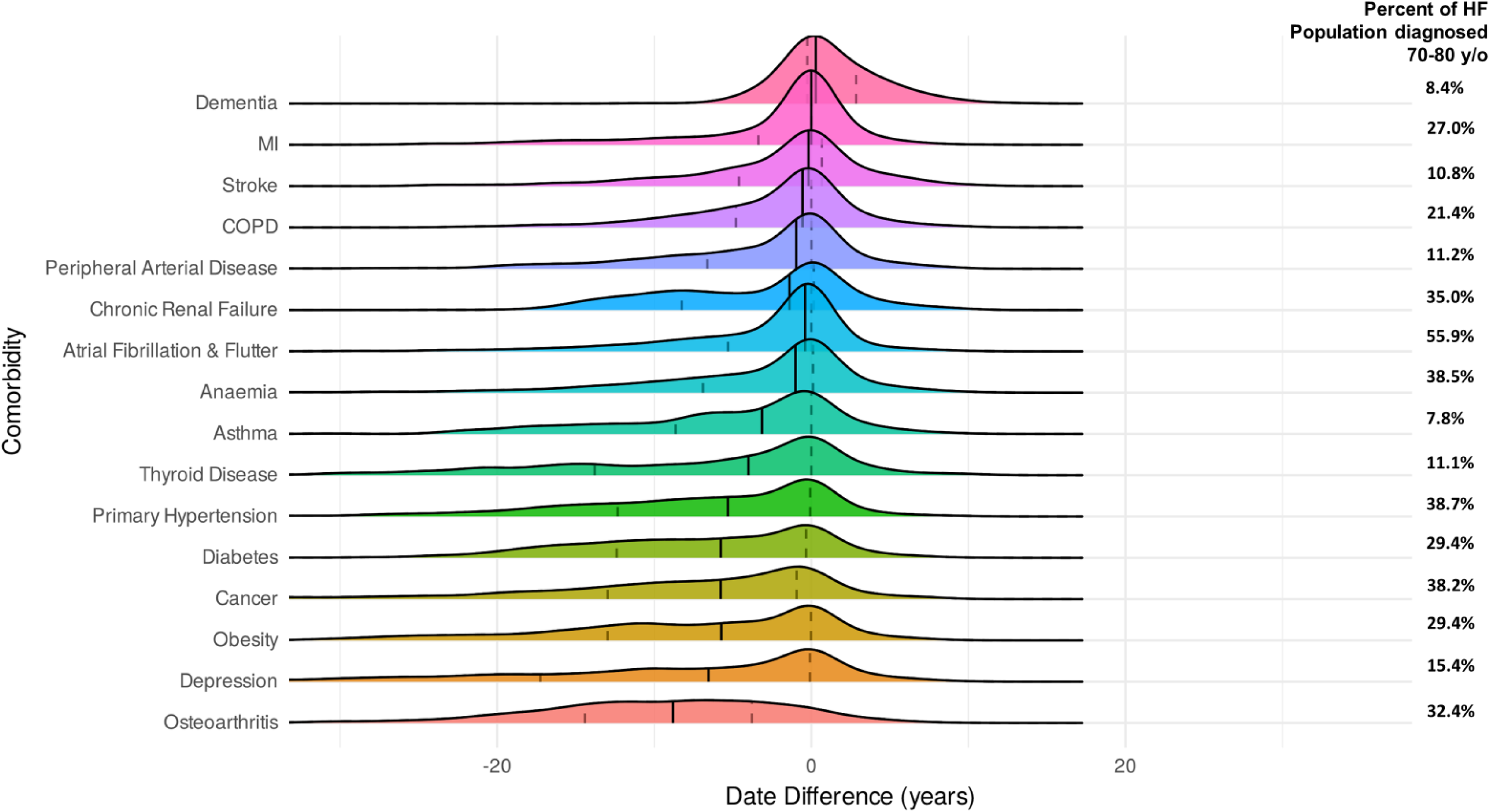
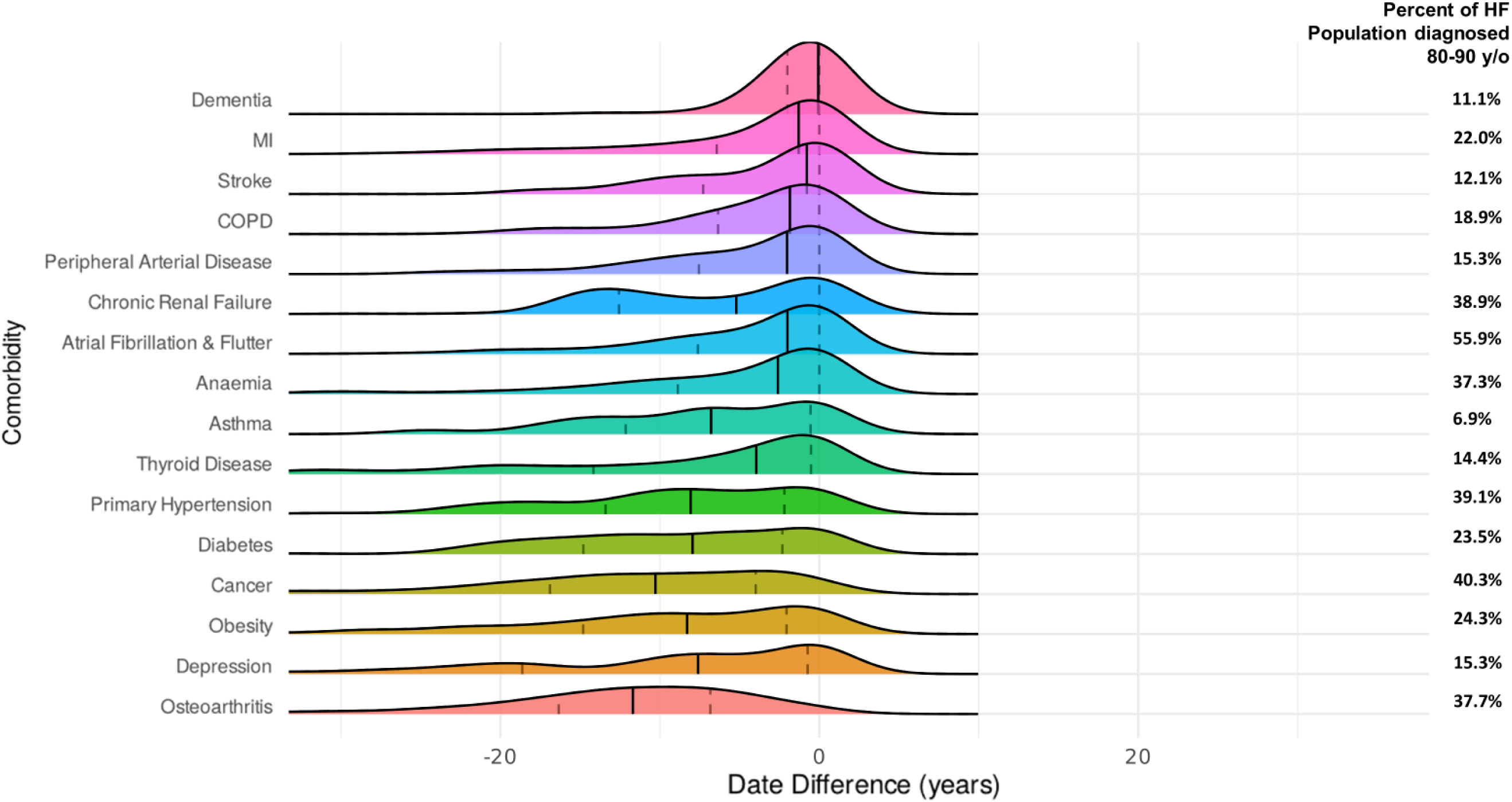
Timing of comorbidity diagnoses in relation to HF diagnosis stratified by age at HF diagnosis. Ridgeline plots illustrating the chronological distribution of comorbidity diagnoses in relation to heart failure (HF) diagnosis for each stratum of age at HF diagnosis. The amplitude of Ridgelines does not relate to comorbidity prevalence, the data for which are noted to the right of the plot for reference. MI - myocardial infarction; COPD – chronic obstructive pulmonary disease.

## Notes

### Competing Interest Statement

SS has received speakers fees honoraria and non-financial support from Astra Zeneca. MFP has received consultancy fees from Astra Zeneca. RC has received speakers fees from Janssen Oncology.

### Funding Statement

This study was funded by the British Heart Foundation (RG/F/22/110076)

### Author Declarations

Research Ethics Service of UK National Health Service approved UK Biobank via application 11/NW/0382.

